# Outcomes of In-hospital Cardiac Arrest: Insights from a Medical Intensive Care Unit

**DOI:** 10.64898/2025.12.28.25343110

**Authors:** Karthik Kailasam, Xiaozhen Han, Xiaofeng Wang, Sudhir Krishnan

## Abstract

**Background:** Critically ill patients admitted to intensive care units (ICU) usually suffer from life-threatening illnesses, and many are hemodynamically unstable. The incidence of cardiac arrest in the ICU is approximately 22 per 1000 admissions, and survival to discharge after in-hospital cardiac arrest (IHCA) is approximately 14%. Variables associated with IHCA survival are poorly understood and the outcomes of cardiopulmonary resuscitation (CPR) in the ICU are poorly reported in the literature. We investigated the characteristics of IHCA and factors that are associated with poor IHCA survival.

**Results:** After adjusting for age, APACHE III score, and initial rhythm, every one-minute increase in CPR duration was associated with 1.161 (95% CI 1.119-1.204; p<0.0001) odds of death during resuscitation and 1.154 (95% CI 1.059-1.258; p<0.0001) odds of death at the time of ICU discharge. Hospital survivors had a lower APACHE III score (Mean=88.3, SD 29.8, IQR 66-106) and acute physiology score (Mean=75, SD 30, IQR 56-94) compared to non-survivors. Hospital survivors were also more likely than non-survivors to have a shockable rhythm at the time of arrest (20% versus 7.5%), shorter average CPR duration (5.4 minutes versus 12.8 minutes), longer length of ICU stay (14 days versus 1.8 days) and longer length of hospital stay (25 days versus 6.1 days).

**Conclusion:** Based on our retrospective analysis, we conclude that the odds of IHCA mortality is directly proportional to the duration of CPR regardless of age, initial rhythm, and severity of underlying illness.

## INTRODUCTION

There are approximately 292,000 in-hospital cardiac arrests (IHCA) in the United States every year, with one in four survivors (1). The factors associated with survival and favorable neurological outcomes following IHCA are poorly understood. Girotra et al showed improvements in IHCA survival to discharge from 13.7% in 2000, up to 22.3% in 2009. Among those survivors, 85% were discharged from the hospital with a favorable neurological outcome (2). Survival following IHCA decreases with increasing age, especially among elderly patients (age >70) (3). Kagawa et al found IHCA patients were often older and sick with underlying co-morbid conditions (4).

Altered metabolic parameters, inadequate organ perfusion, decompensation of comorbid conditions and medical interventions are some of the variables that can precipitate a cardiopulmonary arrest in the ICU. Moskowitz et al found that delay in responding to impending respiratory failure, incorrect diagnosis, and delay in responding to clinical deterioration were some of the preventable targets for cardiac arrest in ICU (5). Approximately 59% of all IHCA occur in the ICU, with only 14% survival during the hospitalization (6).

Variations in the reported incidence of cardiac arrest, the relevance of these findings to the specific healthcare system, and the potential for under-reporting plague our understanding of IHCA and CPR outcomes in the ICU (7). In our retrospective observational study, we attempt to identify patient characteristics of IHCA in the ICU and determine the impact of duration of cardiopulmonary resuscitation (CPR) on survival outcomes. We hypothesize that prolonged CPR in critically ill patients admitted to the ICU is associated with higher mortality among those who survive the initial resuscitation.

## MATERIALS AND METHODS

### Data source and Study population

We conducted an institutional review board-approved, single-center, 5-year retrospective study of all critically ill patients > 18 years of age (N=367) who had an in-hospital cardiac arrest (January 2014 to December 2018). Patients were admitted to the 64-bed adult medical intensive care unit at the Cleveland Clinic. The study was approved by the institutional review board of Cleveland Clinic Foundation (Study No. 19 820). We used REDcap (Research electronic data capture) to create de-identified datasets, removing the 18 HIPAA identifiers, for handling Protected Health Information.

Clinical data were obtained from Epic® electronic medical record. The following data points were extracted and analyzed: Demographics, Acute Physiology and Chronic Health Evaluation (APACHE) III score, Acute Physiology Score (APS), initial rhythm at the time of cardiac arrest, duration of CPR, length of hospital stay, length of ICU stay, survival after ROSC, survival at ICU discharge, survival at hospital discharge, and underlying comorbid medical conditions. Comorbid conditions included in our data collection are diabetes mellitus, chronic kidney disease on dialysis (CKD), chronic obstructive lung disease (COPD), cirrhosis, presence of immunosuppression, sepsis, and malignancy. Acute coronary syndromes leading to cardiac arrest are admitted to our dedicated cardiac ICU and hence the lack of cardiac comorbid conditions like congestive heart failure (CHF) in our study population. APACHE III scores were calculated for each patient from data collected during the first 24 hours of ICU admission.

### Statistical analysis

Chi-square test was used to evaluate the effects of categorical variables (i.e., gender, admission source, ROSC, work shift, co-morbid conditions, and condition on ICU discharge) on the duration of CPR. Wilcoxon rank-sum test was used to examine the effect of continuous variables (age, length of stay, and APACHE III score) on the duration of CPR. The study variables significant at P < .10 in univariate analyses were explored in multivariate regression models. Variables significant at P < .05 were selected in the final regression models. Correlation analysis of the predictors was conducted to avoid multicollinearity in the model of mortality. Association between these variables was expressed as an odds ratio with 95% CI. Statistical analyses were performed using SAS 9.4 software (SAS Institute). The level of statistical significance was set at P < .05 (two-tailed).

## RESULTS

The baseline characteristics of our study population and characteristics of IHCA based on the duration of resuscitation are noted in table 1 and table 2 respectively. The mean age was 61 years (IQR, 51-73) of which 55% were men. The mean duration of resuscitation was 11.5 minutes (IQR, 4-17). Return of spontaneous circulation (ROSC) was obtained in 224 patients (61%) of which 82 survived to ICU discharge (23.4%) and 60 patients (16.3%) survived to hospital discharge. Prolonged CPR (>15 minutes) was associated with poor outcomes (N= 6; 5.5%).

### Survival based on duration of CPR

After adjusting for age, APACHE III score, and initial rhythm; every one-minute increase in CPR duration was associated with 1.161 (95% CI 1.119-1.204; p<0.0001) odds of death during acute resuscitation and 1.154 (95% CI 1.059-1.258; p<0.0001) odds of death at the time of ICU discharge. A Kaplan-Meier plot to demonstrate the survival probability with the duration of resuscitation is shown in figure1.

**Fig 1.**
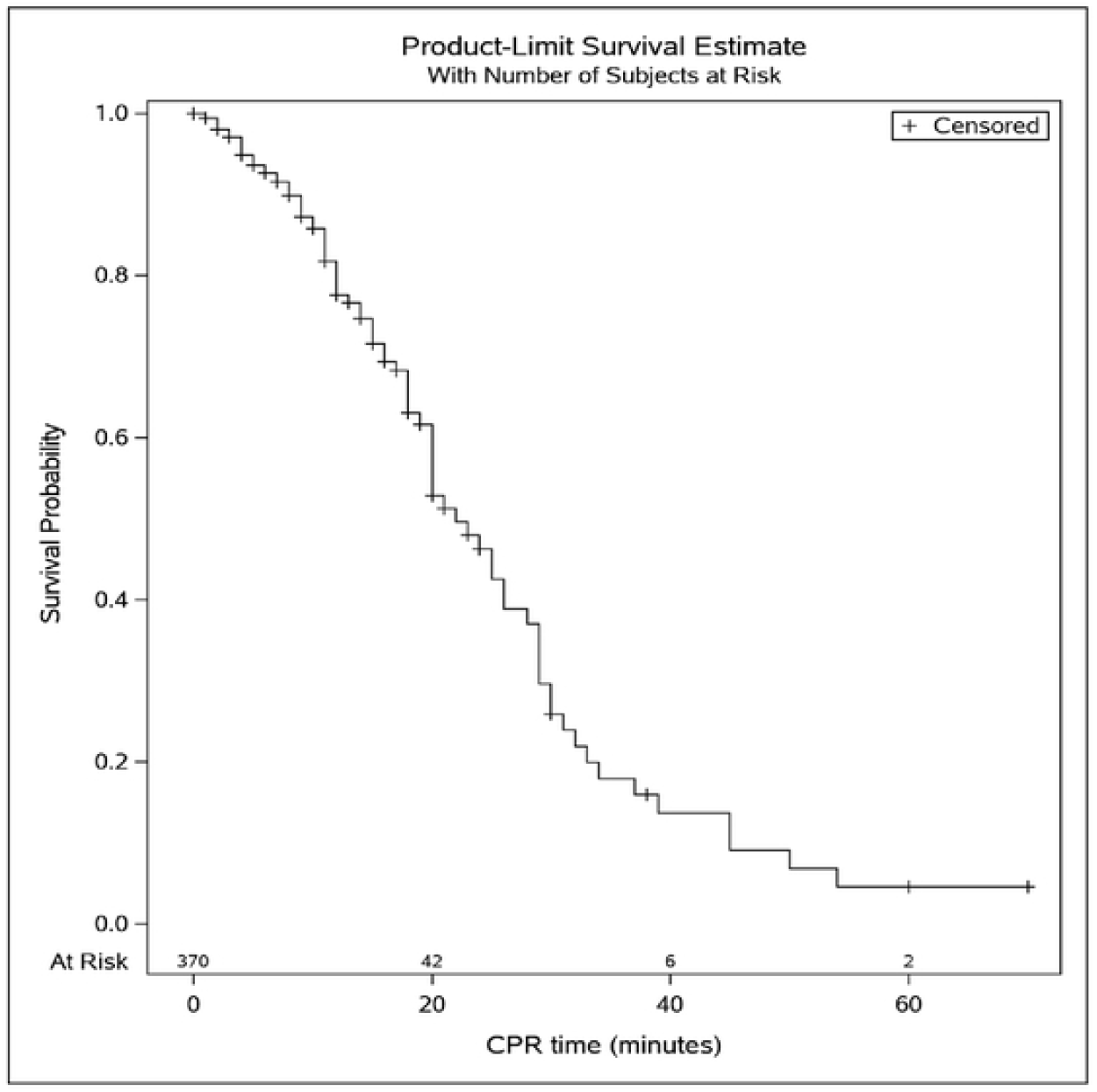
Kaplan-Meier plot for survival probability based on duration of CPR

**Fig 2.**
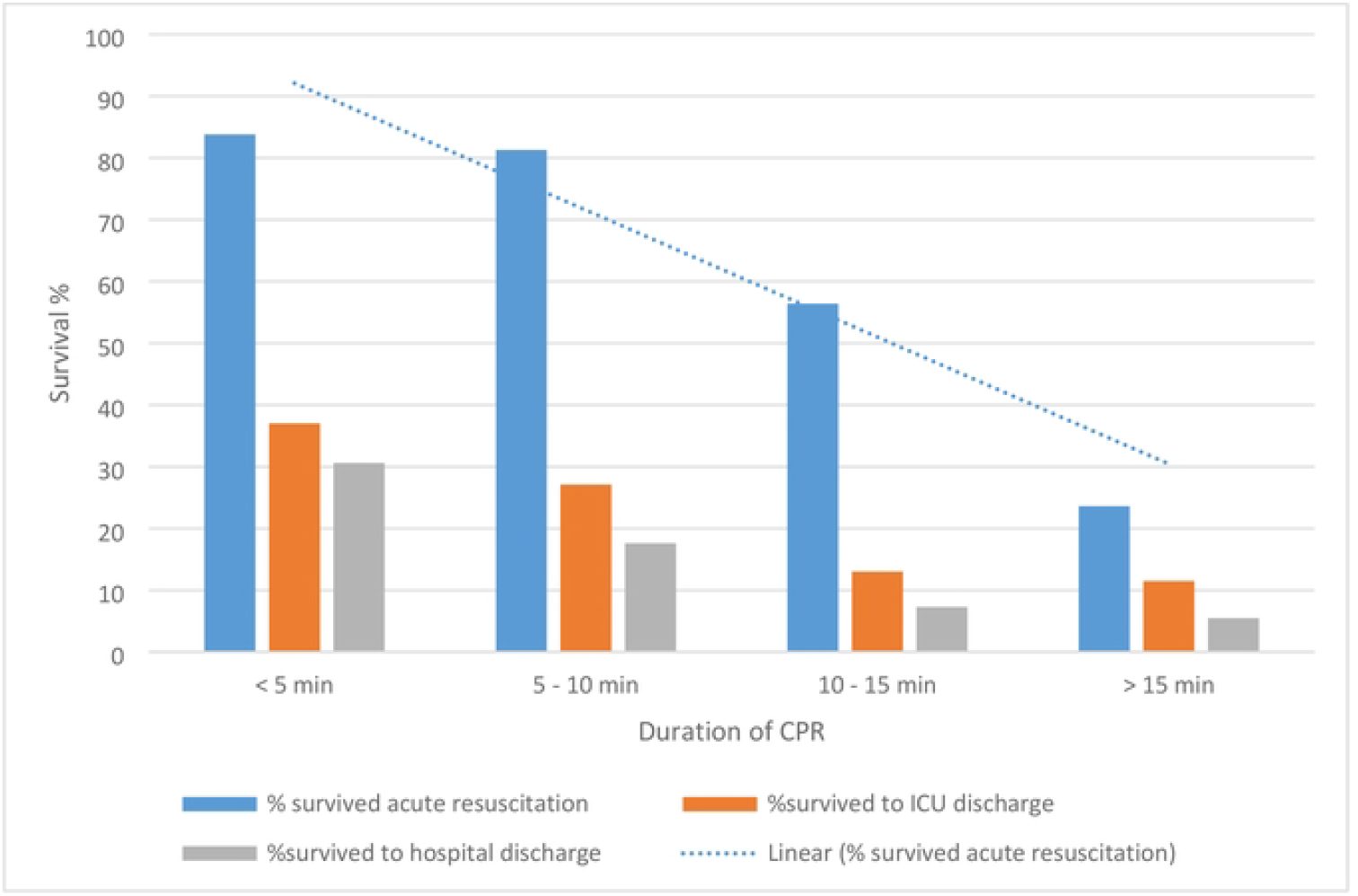
Survival outcomes based on duration of CPR

### Hospital survival

Sixty patients survived to hospital discharge (16.3%). Hospital survivors were marginally younger (mean age of 59; SD 18) of which 55% were men. The APACHE III score (Mean=88.3, SD 29.8, IQR 66-106) and acute physiology score (Mean=75, SD 30, IQR 56-94) were lower compared to non-survivors. Hospital survivors were more likely than non-survivors to have a shockable rhythm at the time of arrest (20% vs 7.5%), shorter average CPR duration (5.4 minutes versus 12.8 minutes), a longer length of ICU stay (14 days versus 1.8 days) and longer length of hospital stay (25 days versus 6.1 days). Characteristic features of hospital survivors and non-survivors are listed in table 3.

## DISCUSSION

In our study, we report ICU-IHCA survival of 16.3% at hospital discharge in comparison to previously reported 14% survival to discharge after IHCA in ICU patients over the years 2003-2010 (6). Improved survival rate in our study population is likely related to continued advancements in resuscitation methods and clinical care over the last decade (8,9).

In our analysis, patients with CPR lasting longer than 15 minutes were relatively younger with a mean age of 59 (SD 15.9), compared to patients with CPR lasting less than 5 minutes with a mean age of 64 (SD 14.9). Khan et al reported that younger age (18-40 vs >65 years; odds ratio [OR] 1.81; 95% CI 1.69 to 1.95; *P*<0.001) and female sex (OR 1.05; 95% CI 1.02 to 1.09; *P*=0.005) were associated with longer duration of resuscitation in patients who did not obtain ROSC after cardiac arrest (10). It is assumed that this observed difference could be related to a perception among health care workers that younger patients have a higher probability of survival and merit a longer duration of resuscitation (9). In contrast to our study results, a nationwide Swiss data analysis by Amacher et al reported women were less frequently admitted to ICUs, received fewer advanced treatments, and had a higher risk of ICU mortality when compared to men (11).

Patients with chronic illnesses such as CHF, CKD, COPD, malignancy, and cirrhosis who had IHCA were found to have significantly higher mortality in our analysis. Stapleton et al reported increased risk for death after IHCA among older adults with severe COPD (Hazard ratio 1.39; 95%CI 1.32-1.48; p<0.001), severe CHF (HR 1.81; 95% CI 1.75-1.87; p<0.001), severe CKD (HR 1.58; 95% CI 1.52-1.64; p<0.001), severe cirrhosis (HR 1.69; 95% CI 1.40-2.05; p<0.001) and malignancy (HR 1.70; 95% CI 1.58-1.84; p<0.001) when compared to patients without those co-morbid conditions (12). Our patients were acutely ill with a mean APACHE III score of 102.3. In our study population, 17.4 % were on chronic dialysis, 32.5 % had diabetes, 17.4 % had severe COPD, 11.7% had cirrhosis and 30.8 % were immunosuppressed (table1).

Furthermore, 27% of our IHCA patients had sepsis with a mortality rate of 87% at hospital discharge. This is similar to IHCA mortality in patients with severe sepsis reported by Koivikko et al (13). Champigneulle et al reported a six-month survival rate of 14% among cancer patients admitted to the Intensive care unit (ICU) after a cardiac arrest (14). In our cohort, 25% had an underlying malignant condition (leukemia, lymphoma, or solid tumor with metastasis) with a mortality rate of 93% at hospital discharge.

In our study, we noticed an increase in mortality proportional to the duration of resuscitation. Fernando et al in their meta-analysis of 23 cohort studies reported lower odds of survival (OR 0.12; 95% CI 0.07 to 0.19; p<0.001) in patients with CPR longer than 15 minutes (15). Radeschi et al studied the outcomes of IHCA in Italy and reported the median duration of CPR for survivors was 8 minutes and 26 minutes for non-survivors (16). Kantamineni et al and Rafati et al also reported better outcomes when CPR lasted less than 10 minutes (17,18). In our study, the mean duration of CPR was 5.2 minutes (IQR 2-6) for patients who survived to ICU discharge; and 16.4 minutes (IQR 7-24) for patients who died during acute resuscitation.

Prolonged resuscitation is associated with various traumatic and non-traumatic complications (19). There is no clear definition for prolonged CPR as the ideal duration of resuscitation is unknown. Loisa et al reported that approximately 14% of IHCA are terminated early due to presumed futility (20). American Heart Association’s cardiac arrest guidelines do not have any specific recommendations on termination of CPR.

Lately, clinical decision rules have been developed to assist physicians in terminating potentially futile resuscitation attempts (21). Val Walraven et al proposed a model (the UN10 rule) for ‘futile CPR’ incorporating three intra-arrest variables i) unwitnessed arrest ii) non-shockable rhythm and iii) ROSC not obtained in 10 minutes (22,23). Petek et al reported that among patients who meet the UN10 criteria for futile resuscitation, only 6.3% survived to discharge and 4.8% survived with favorable neurological function (23). Our cohort had witnessed cardiac arrest in a monitored hospitalized setting, however, among non-survivors, 93% had a non-shockable rhythm and an average CPR duration of 12.8 minutes.

### Strengths and limitations

The key strengths of our study include the seminal nature of attempting to explore outcomes of IHCA in the ICU with a relatively large sample size. The study may be representative of IHCA ICU outcomes in comparable critical care setting. We acknowledge the limitations of our study. First, our retrospective study was conducted in a single large academic center involved in the care of complex patients that might be reflective of patient demographics at other institutions. Acute coronary syndrome, a common cause for IHCA is not included in our study population since they were admitted to our dedicated cardiac ICU (24). Hence, the lack survival outcomes based on cardiac comorbid conditions in our study population.

The GO-FAR score (Good outcome following resuscitation) classifies patients based on the likelihood of survival with good neurological outcomes on discharge following IHCA (21). We were not able to incorporate the GO-FAR score in our model due to the lack of all 13 variables which are required for the calculation. We do not have data on neurological recovery after IHCA but only overall functional status at the time of discharge. Due to limitations in sample size, we were unable to perform a multivariate regression analysis to exclude all confounding factors which could influence survival outcomes.

## Conclusion

Based on our study findings, the mortality after IHCA increases with the increase in the duration of CPR regardless of age, initial rhythm, and severity of underlying illness. Further studies using clinical decision rules like GO-FAR score might assist the healthcare team in selecting appropriate patients who might benefit from prolonged resuscitation.

## Data Availability

If the data are all contained within the manuscript and/or Supporting Information files, enter the following: All relevant data are within the manuscript and its Supporting Information files. Due to institutional policies we cannot disclose protected health information

## References

1. Holmberg MJ, Ross CE, Fitzmaurice GM, Chan PS, Duval-Arnould J, Grossestreuer AV, et al. Annual Incidence of Adult and Pediatric In-Hospital Cardiac Arrest in the United States. Circ Cardiovasc Qual Outcomes. 2019/07/09 ed. 2019 July;12(7):e005580.

2. Girotra S, Nallamothu BK, Spertus JA, Li Y, Krumholz HM, Chan PS. Trends in Survival after In-Hospital Cardiac Arrest. N Engl J Med. 2012 Nov 15;367(20):1912–20.

3. van Gijn MS, Frijns D, van de Glind EM, C van Munster B, Hamaker ME. The chance of survival and the functional outcome after in-hospital cardiopulmonary resuscitation in older people: a systematic review. Age Ageing. 2014/04/22 ed. 2014 July;43(4):456–63.

4. Kagawa E, Inoue I, Kawagoe T, Ishihara M, Shimatani Y, Kurisu S, et al. Assessment of outcomes and differences between in- and out-of-hospital cardiac arrest patients treated with cardiopulmonary resuscitation using extracorporeal life support. Resuscitation. 2010/06/02 ed. 2010 Aug;81(8):968– 73.

5. Moskowitz A, Berg KM, Cocchi MN, Chase M, Yang JX, Sarge J, et al. Cardiac arrest in the intensive care unit: An assessment of preventability. Resuscitation. 2019/09/12 ed. 12;145:15–20.

6. Perman SM, Stanton E, Soar J, Berg RA, Donnino MW, Mikkelsen ME, et al. Location of In-Hospital Cardiac Arrest in the United States-Variability in Event Rate and Outcomes. J Am Heart Assoc [Internet]. 2016/09/29 ed. 9;5(10). Available from: https://www.ncbi.nlm.nih.gov/pubmed/27688235

7. Armstrong RA, Kane C, Oglesby F, Barnard K, Soar J, Thomas M. The incidence of cardiac arrest in the intensive care unit: A systematic review and meta-analysis. J Intensive Care Soc. 2019 May;20(2):144–54.

8. Thompson LE, Chan PS, Tang F, Nallamothu BK, Girotra S, Perman SM, et al. Long-Term Survival Trends of Medicare Patients After In-Hospital Cardiac Arrest: Insights from Get With The Guidelines-Resuscitation. Resuscitation. 2017/11/02 ed. 2;123:58–64.

9. Wiberg S, Holmberg MJ, Donnino MW, Kjaergaard J, Hassager C, Witten L, et al. Age-dependent trends in survival after adult in-hospital cardiac arrest. Resuscitation [Internet]. 2020/04/01 ed. 2020 Apr; Available from: https://www.ncbi.nlm.nih.gov/pubmed/32246986

10. Khan AM, Kirkpatrick JN, Yang L, Groeneveld PW, Nadkarni VM, Merchant RM, et al. Age, sex, and hospital factors are associated with the duration of cardiopulmonary resuscitation in hospitalized patients who do not experience sustained return of spontaneous circulation. J Am Heart Assoc. 2014 Dec;3(6):e001044.

11. Amacher SA, Zimmermann T, Gebert P, Grzonka P, Berger S, Lohri M, et al. Sex disparities in ICU care and outcomes after cardiac arrest: a Swiss nationwide analysis. Crit Care. 2025 Jan 23;29(1):42.

12. Stapleton RD, Ehlenbach WJ, Deyo RA, Curtis JR. Long-term outcomes after in-hospital CPR in older adults with chronic illness. Chest. 2014 Nov;146(5):1214–25.

13. Koivikko P, Arola O, Inkinen O, Tallgren M. One-Year Survival after Inhospital Cardiac Arrest-Does Prearrest Sepsis Matter? Shock. 7;50(1):38–43.

14. Champigneulle B, Merceron S, Lemiale V, Geri G, Mokart D, Bruneel F, et al. What is the outcome of cancer patients admitted to the ICU after cardiac arrest? Results from a multicenter study. Resuscitation. 2015/04/24 ed. 2015 July;92:38–44.

15. Fernando SM, Tran A, Cheng W, Rochwerg B, Taljaard M, Vaillancourt C, et al. Pre-arrest and intra-arrest prognostic factors associated with survival after in-hospital cardiac arrest: systematic review and meta-analysis. BMJ. 2019/12/04 ed. 2019 Dec;367:6373.

16. Radeschi G, Mina A, Berta G, Fassiola A, Roasio A, Urso F, et al. Incidence and outcome of in-hospital cardiac arrest in Italy: a multicentre observational study in the Piedmont Region. Resuscitation. 2017/06/24 ed. 10;119:48–55.

17. Kantamineni P, Emani V, Saini A, Rai H, Duggal A. Cardiopulmonary resuscitation in the hospitalized patient: impact of system-based variables on outcomes in cardiac arrest. Am J Med Sci. 2014 Nov;348(5):377–81.

18. Rafati H, Saghafi A, Saghafinia M, Panahi F, Hoseinpour M. Survival after In-Hospital Cardiopulmonary Resuscitation in a Major Referral Center during 2001-2008. Iran J Med Sci. 2011 Mar;36(1):50–3.

19. Youness H, Al Halabi T, Hussein H, Awab A, Jones K, Keddissi J. Review and Outcome of Prolonged Cardiopulmonary Resuscitation. Crit Care Res Pr. 2016/01/14 ed. 2016;2016:7384649.

20. Loisa E, Setälä P, Hoppu S, Tirkkonen J. Early termination of resuscitation in in-hospital cardiac arrest and impact to the outcome calculations. Acta Anaesthesiol Scand. 2019/07/21 ed. 10;63(9):1239– 45.

21. Thai TN, Ebell MH. Prospective validation of the Good Outcome Following Attempted Resuscitation (GO-FAR) score for in-hospital cardiac arrest prognosis. Resuscitation. 2019/05/09 ed. 7;140:2–8.

22. van Walraven C, Forster AJ, Parish DC, Dane FC, Chandra KM, Durham MD, et al. Validation of a clinical decision aid to discontinue in-hospital cardiac arrest resuscitations. JAMA. 2001 Mar;285(12):1602–6.

23. Petek BJ, Bennett DN, Ngo C, Chan PS, Nallamothu BK, Bradley SM, et al. Reexamination of the UN10 Rule to Discontinue Resuscitation During In-Hospital Cardiac Arrest. JAMA Netw Open. 2019/05/03 ed. 5;2(5):e194941.

24. Zhang Y, Yu Y, Qing P, Liu X, Ding Y, Wang J, et al. In-hospital cardiac arrest characteristics, causes and outcomes in patients with cardiovascular disease across different departments: a retrospective study. BMC Cardiovasc Disord. 2024 Sept 6;24:475.

